# The quality of vital signs measurements in electronic medical records varies by hospital, specialty, and patient demographics

**DOI:** 10.1101/2022.01.19.22269544

**Authors:** Niall Jackson, Jessica Woods, Peter Watkinson, Andrew Brent, Tim EA Peto, A Sarah Walker, David W Eyre

## Abstract

**Objective:** To assess the frequency of digit preference in recording of vital signs in electronic healthcare records (EHRs) and associated patient and hospital factors.

**Study Design and Setting:** We used EHR data from Oxford University Hospitals, UK, between 01-January-2016 and 30-June-2019 and multivariable logistic regression to investigate associations between temperature readings of 36.0°C or systolic and diastolic blood pressure (SBP/DBP) readings both ending in zero and patient age, sex, ethnicity, deprivation, comorbidities, calendar time, hour of day, days into admission, hospital, day of week and speciality.

**Results:** In 4,305,914 records from 143,352 patients, there was an excess of temperature readings of 36.0°C (15.1%, 649,976/4,305,914), compared to an expected 4.9% from the underlying distribution. 2.2% (95,215) BP readings had a SBP and DBP both ending in zero vs. 1% expected by chance. Digit preference was more common in older and male patients, as length of stay increased, following a previous normal set of vital signs and typically more common in medical vs. surgical specialities. Differences were seen between hospitals, however, digit preference reduced over calendar time.

**Conclusion:** Vital signs may not always be accurately documented. Allowances and adjustments may be needed in observational analyses using these factors as outcomes or exposures.

**What’s New?:** *Key findings:*

▪ Digit preference in the recording of vital signs in electronic healthcare records is common, affecting approximately 10% of temperature measurements and 1% of blood pressure recordings in a large UK teaching hospital group
▪ These findings were obtained in hospitals using a semi-automated data capture system that required manual re-entry of vital signs into a tablet computer prior to automated upload to electronic patient records
▪ Digit preference was associated with patient characteristics and was more common in older and male patients, as length of stay increased and following previously normal vital signs
▪ Digit preference varied between hospitals, but decreased over time
▪ Digit preference was generally more common in medical compared to surgical specialties

*What this adds to what is known:*

▪ Most previous studies of data quality in electronic patient records have focused on the accuracy of coding
▪ This study focuses on the accuracy of numeric values in patient records, and also adds new data on patient and hospital factors associated with the accuracy of values in electronic patient records

*Implications:*

▪ Clinicians and researchers need to be aware that vital signs may not always be accurately documented
▪ Appropriate allowances and adjustments for digit preference should be considered in observational analyses using these factors as outcomes or exposures.
▪ Further work is required to understand the mechanisms behind values preference on a systems, patient and clinician level

## Introduction

Electronic healthcare records (EHR) have been widely adopted across different healthcare settings globally and have become an integral part of health infrastructure: saving time, improving communication and record keeping, and supporting learning.^1^ As the scale and breadth of EHR data increases, so does its ability to fulfil secondary functions including quality improvement, product development, and research, contingent on appropriate regulation and transparency.^2^ Example applications of EHR data include population-level epidemiological studies,^3–5^ machine learning-based diagnostic assistants for clinicians,^6^ screening for child maltreatment and family violence,^7^ and detecting and tracking infectious disease outbreaks.^8,9^

However, conclusions rely on the reliability and accuracy of EHR data, which is not guaranteed.^10,11^ Indeed, the use of EHR data beyond its original purpose (clinical care and billing) raises specific challenges. Data collection in the clinical environment is imperfect^12^ and often incomplete;^13^ it may lack comparability or reproducibility^14^ or even simply be wrong.^15,16^ Attempts to quantify or evaluate EHR data quality are limited, and even fewer have investigated causes for variability in quality.^17^ Most studies have focused on checking the accuracy of clinical and diagnostic codes rather than numerical observations. In one notable exception, evaluating the quality of vital sign data across multiple hospitals and EHR systems, there was a skew of completeness and correctness in favour of arriving patients and higher fidelity in wholly EHR based systems compared to a combination of paper and EHR.^18^

Digit preference in vital sign measurement is well recognized in healthcare. Terminal digit preference for multiples of ten in blood pressure recordings has been shown to extremely common,^19,20^ introduce systemic bias potentially effecting mortality^21^ and produce inaccurate epidemiological results.^22^ This phenomenon has been observed in other vital sign measurements such as respiratory rate, with attempts to rectify inaccuracy through continuous, automated monitoring.^23^

In this study we investigate observations of vital signs gathered over 3.5 years from inpatients at a large UK teaching hospital group. We assess the frequency of recordings with preferences for a specific value (e.g., multiples of ten for blood pressure or temperature readings of 36.0°C) as a marker of sub-optimal data quality and establish associated patient and hospital factors.

## Methods

### Setting

We conducted a retrospective observational study at Oxford University Hospitals NHS Foundation Trust (OUH). OUH consists of four teaching hospitals with a total of 1000 beds: Hospital A (providing acute care, trauma and neurosurgery services); Hospital B (elective cancer surgery, transplant, haematology, oncology); Hospital C (district hospital, acute medical services) and Hospital D (elective orthopaedics). OUH acts as a tertiary referral centre for the surrounding region, generating approximately 1 million patient contacts a year, serving a population of around 655,000.

### Data

Deidentified data were obtained from the Infections in Oxfordshire Research Database which has approvals from the National Research Ethics Service South Central – Oxford C Research Ethics Committee (19/SC/0403), the Health Research Authority and the national Confidentiality Advisory Group (19/CAG/0144). We used individual observations of vital signs conducted in OUH for adult patients (≥18y) between 01-January-2016 and 30-June-2019. The vital signs observed, with dates and times of collection, included respiratory rate (RR), heart rate (HR), tympanic temperature, systolic and diastolic blood pressure (SBP and DBP).

Observations were collected using a semi-automated vital sign observation system. HR, SBP and DBP were collected using an observation machine, RR manually timed, and temperature with a separate tympanic thermometer. All observations were then manually transcribed into a tablet computer attached to the same stand, which uploaded results into the EHR. Oxygen saturations, supplemental oxygen and alertness (alert, responsive to voice, pain or unresponsive, AVPU) are also all recorded. However, these measurements are not considered further here as the dynamic range of oxygen saturation measurements or number of categories for AVPU scoring were insufficient to distinguish digit preference from the underlying distribution of the data.

Additional data were obtained: hospital-level data (hospital and ward where the observation was made, the specialty managing the patient); and patient data (age, sex, ethnic group, index of multiple deprivation (IMD) score at home address, Charlson comorbidity score).

### Statistical analysis

We defined preferences for a specific value or digit for each vital sign based on the observed distribution of values. Temperatures of 36.0°C, blood pressure (BP) with both SBP and DBP recorded as multiples of ten, RRs of 16 or 18, and HRs as multiples of ten were all more common than would be expected by chance.

Temperature had the clearest value preference signal; associated factors were investigated using multivariable logistic regression. Analyses were restricted to patients with complete data, and to complete vital sign sets (i.e. all of temperature, HR, RR, SBP, DBP recorded). We used natural cubic splines to account for non-linear relationships for continuous variables (allowing up to five default placed knots, selecting the final number of knots by minimising the Bayesian Information Criterion, BIC). To avoid undue influence of outlying values, continuous variables were truncated at the 1st and 99th percentiles. Pairwise interactions between model main effects were included where this improved model fit based on BIC. We used clustered robust standard errors to account for repeated measurements obtained from the same patient.

To investigate if associations were specific to temperature or applied to vital signs more widely, we also refitted the same model (i.e., with the same spline terms and interactions) with BP digit preference as the outcome. For RR the most common value preferences were also the most common normal values precluding separate investigation of value preference. Value preferences were less frequent for HR and not investigated further.

We also investigated if the presence of abnormal previous temperature or BP readings affected subsequent digit preference. Temperatures of ≤35.5°C or ≥37.5°C and SBP readings of >160 or <90 mmHg or DBP readings of >100 or <60 mmHg were arbitrarily considered abnormal. For each observation with a prior observation from the same patient within ≤36 hours, we selected the most recent prior observation for comparison. We then refitted the regression models above including if the prior temperature or BP reading had been abnormal as a covariate.

Analyses were conducted using R, version 4.1.

## Results

Between 01-January-2016 and 30-June-2019, a total of 5,799,224 sets of vital signs were recorded. For 482,257 (8.3%) records no deprivation score was documented, and these records were excluded. Complete data were available for all other hospital/patient variables. Within the 5,316,967 vital signs sets included, 758,955 (14.3%) did not include temperature, 473,433 (8.9%) were missing SBP and/or DBP, 535,534 (10.1%) missing RR, and 450,899 (8.5%) missing HR recordings. Restricting to complete sets of vital signs left 4,305,914 (81.0%) records in the final dataset from 143,352 patients. The median (IQR) patient age was 60 (42-75) years, 74,726 (52.1%) were female, and 105,751 (73.8%) were of white and 30,272 (21.1%) of unstated or unknown ethnicity. The most common specialties recording vital signs were general surgery (861,807, 20.0%), trauma and orthopaedics (640,259, 14.9%) and acute and emergency medicine (619,636, 14.4%).

### Prevalence of value preferences in vital signs readings

Compared with the overall distribution of temperature values, there was an excess of temperature readings of 36.0°C (Figure 1), which accounted for 15.1% (649,976/4,305,914) of values. This compared to an expected 4.9% (Figure S1) from the underlying distribution, i.e., approximately 10% of all temperature recordings were likely inappropriately recorded as 36.0°C. Approximately 1% of all BP readings would be expected to have both a SBP and DBP ending in zero, however 2.2% (95,215) of readings showed this pattern. Digit preference was less common for HR readings, with 12.1% (522,101) ending in zero compared to an expected 10%. Although 51.4% (2,213,322) of RR readings were 16 or 18 reflecting a likely excess of these values, RR was not studied further as it was difficult to distinguish which associations reflected normal physiology and which digit preferences.

**Figure 1.**
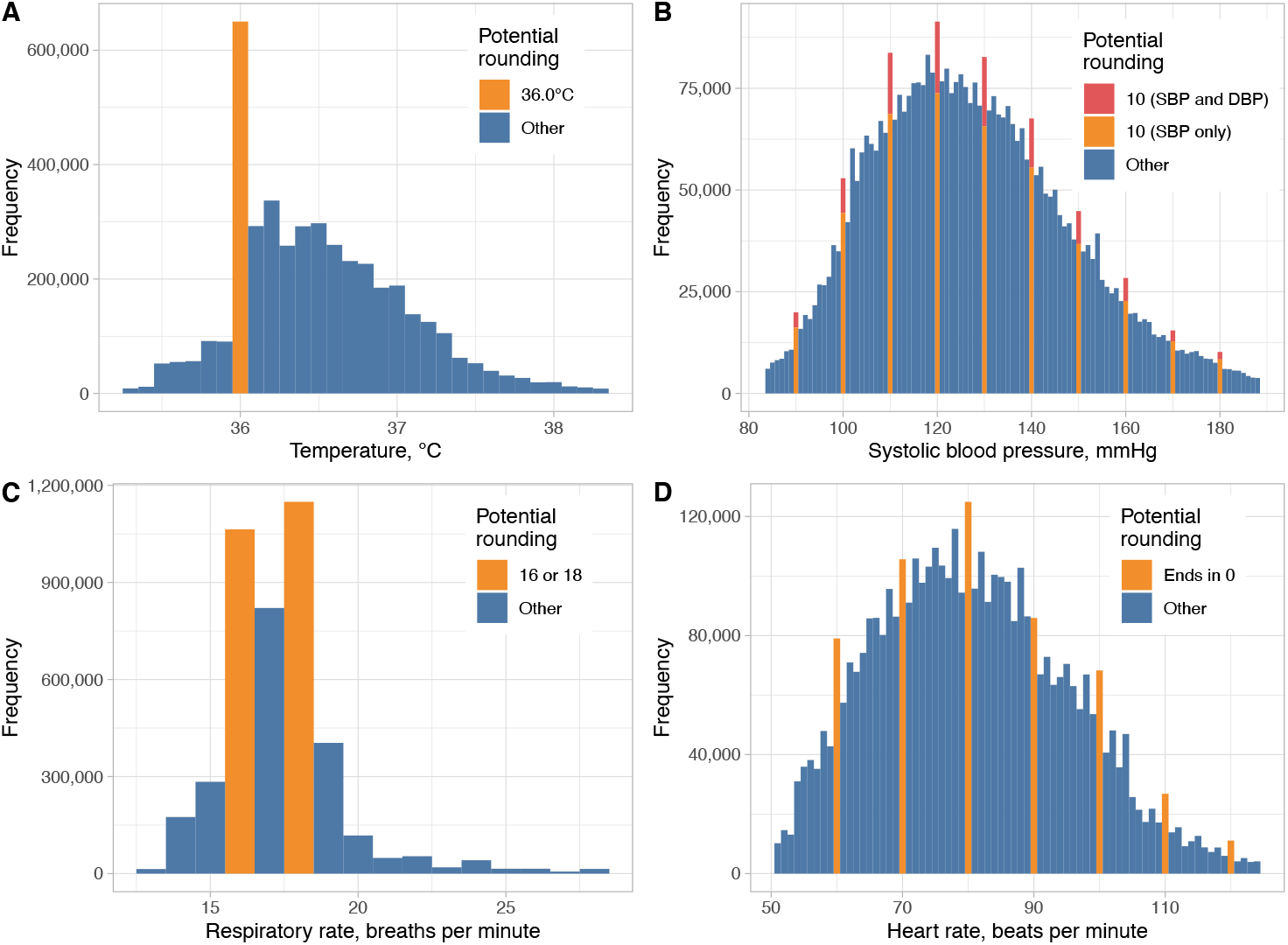
Observed distribution of temperature, systolic blood pressure, respiratory rate and heart rate recordings (n=4,305,914). Readings showing potential value preferences are shown in orange/red. SBP, systolic blood pressure; DBP, diastolic blood pressure. Values below the 1^st^ percentile or above the 99^th^ percentile are omitted for visualisation purposes.

### Value preference associations with patient demographics

Temperature was independently more likely to be recorded as 36.0°C with increasing age above 50 years and BP most likely to be recorded with SBP and DBP both ending in zero for those above 80 years (Figure 2A). Male patients were more likely than female patients to have readings with value preferences for both temperature (adjusted odds ratio, aOR=1.10 [95%CI 1.09-1.12]) and BP (1.04 [1.00-1.08]) (Table 1). Temperature value preference was slightly less common in patients from less deprived areas (aOR per 10 unit change in deprivation percentile=0.99 [0.99-0.99, higher percentiles are less deprived]), but with no evidence of a difference in BP value preference with variation in deprivation. There was no evidence for consistent differences in value preference by ethnicity, however temperatures of 36.0°C were more commonly recorded in patients of Asian ethnicity (aOR vs. white=1.07 [1.02-1.12]) and BPs ending in zero were more common in those of unstated or unknown ethnicity (aOR vs. white=1.12 [1.07-1.18]). Patients with higher Charlson scores were more likely to have recorded temperatures of 36.0°C (aOR per 5 unit increase=1.02 [1.02-1.02]), but less likely to have BP ending in zero (aOR per 5 unit increase=0.93 [0.92-0.94]).

**Table 1.**
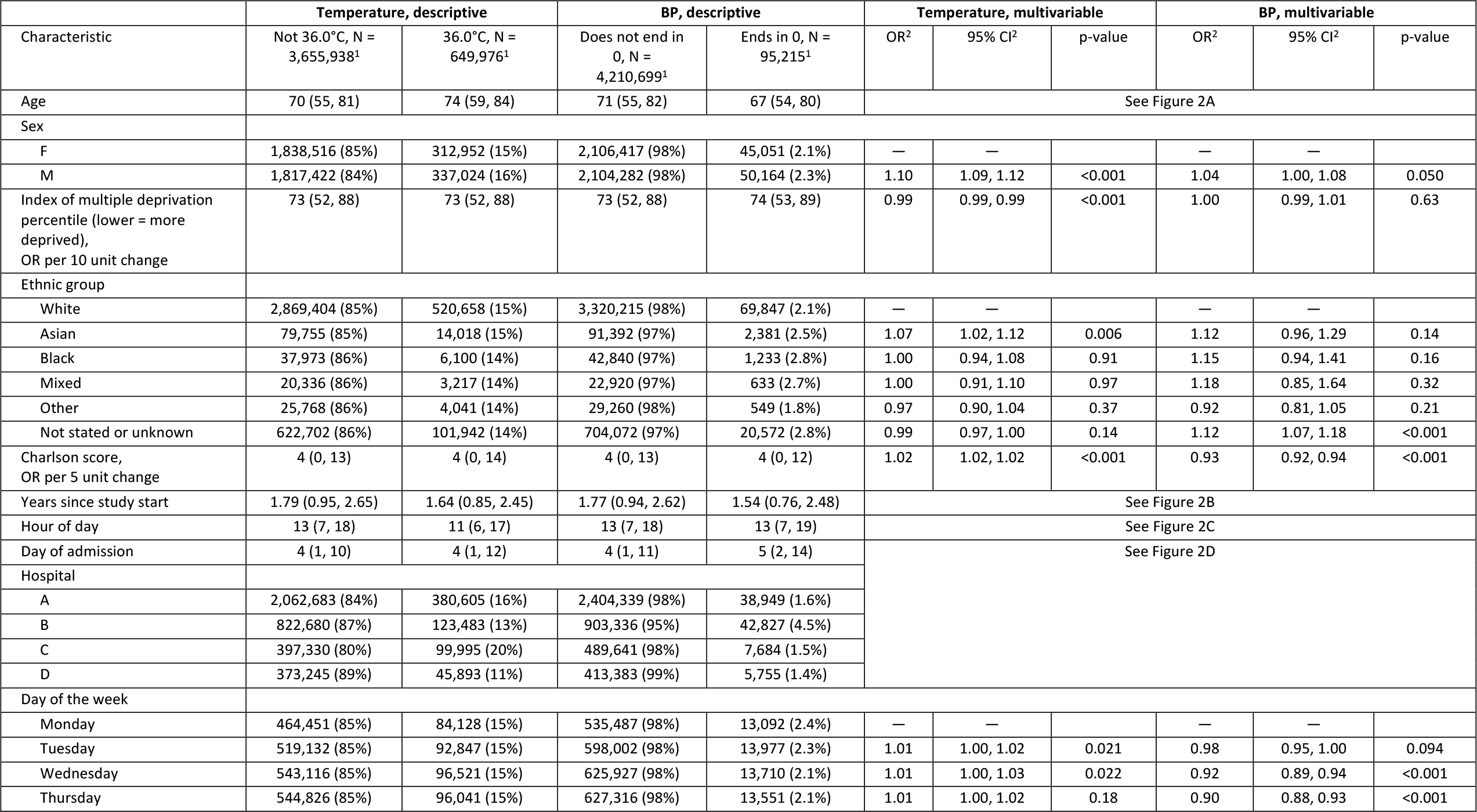
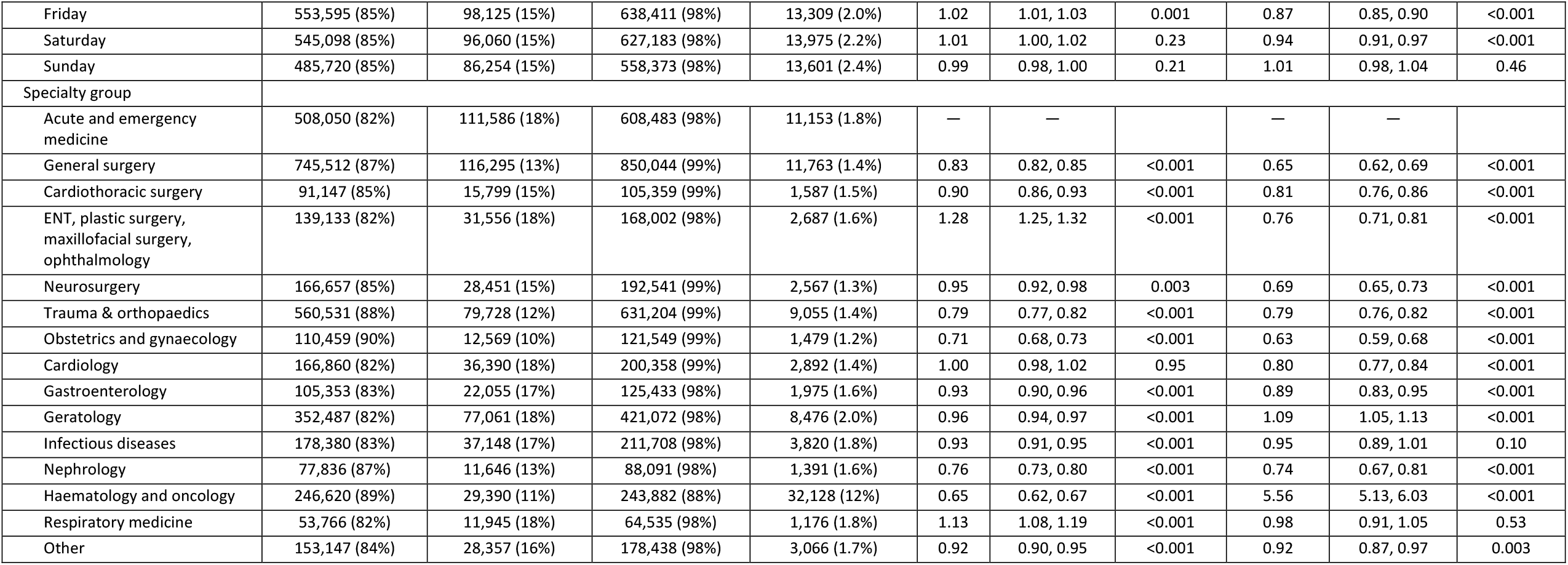
Relationship between temperature readings of 36.0°C and blood pressure readings ending in zero and associated factors. Blood pressure readings where both the systolic and diastolic blood pressure end in zero are shown. Descriptive data for patient factors (age, sex, ethnicity, Index of multiple deprivation, Charlson score), are summarised per observation, rather than per patient. Therefore, for example as older patients had longer hospital stays and more observations the median age is higher than the median age on a per patient basis. ENT, ear, nose and throat surgery; BP, blood pressure. ^1^Median (IQR); n (%). ^2^OR = Odds Ratio, CI = Confidence Interval.

**Figure 2.**
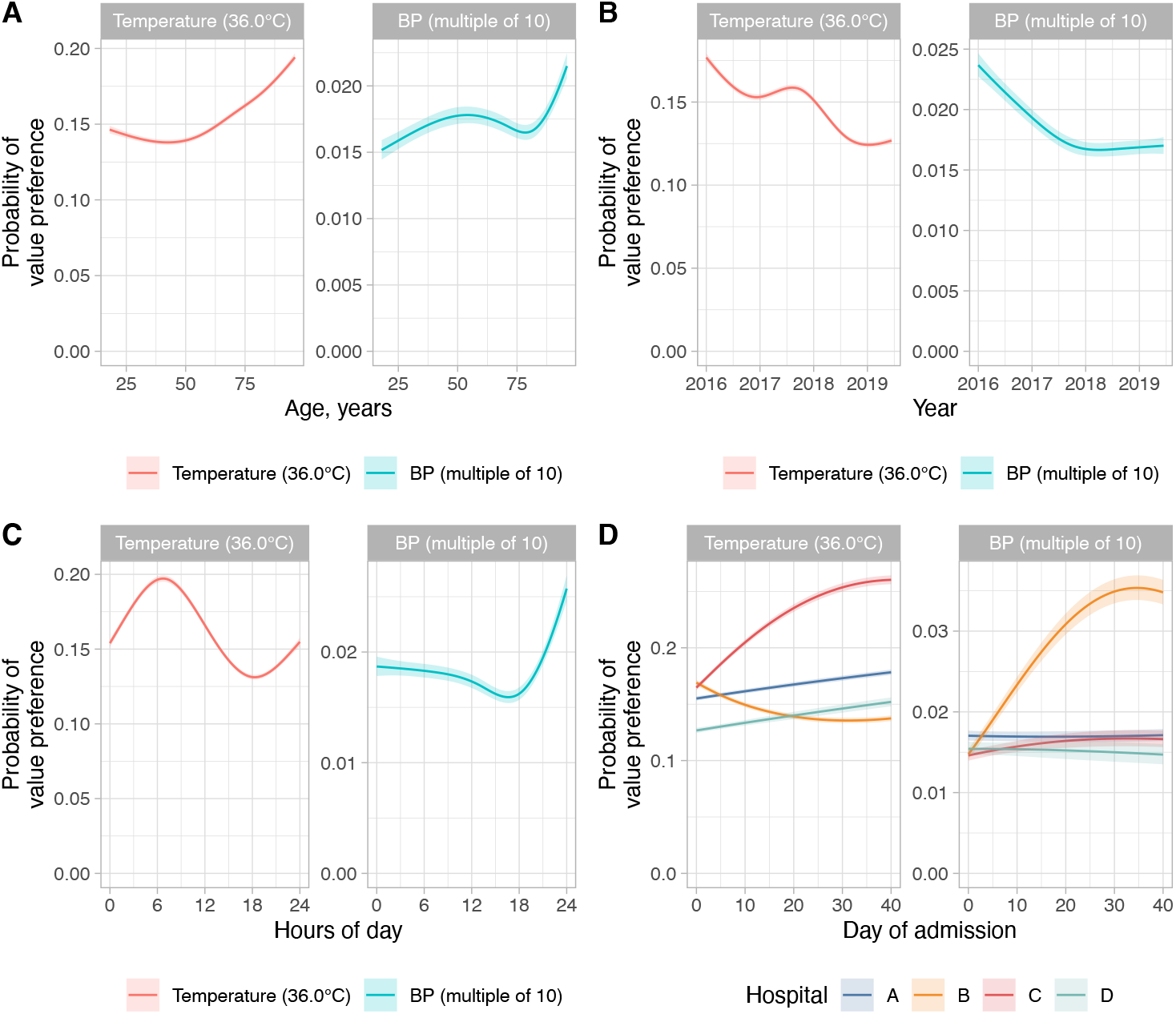
Non-linear associations between factors and temperature readings of 36.0°C and blood pressure readings ending in zero. Model predictions from a multivariable model are shown with all other factors set to the reference category or median value as shown in Table 1. The shaded ribbon indicates the 95% confidence interval. BP, blood pressure.

### Changes over calendar time, hour of day, and by hospital and time in admission

The frequency of temperature and BP measurements with value preferences decreased during the study (Figure 2B), e.g., for temperature from an estimated 17.7% at the start of 2016 to 12.5% at the start of 2019. Temperatures were most likely to be recorded as 36.0°C at around 6am, i.e., at the first routine set of observations performed per day in most patients, whereas BP was most likely to end in zero during the evening (Figure 2C).

Differences were also seen between hospitals in temperature value preferences; at the time of admission this was more common in hospitals A-C, and considerably more common in long stay patients at hospital C. There were small changes in temperature value preferences by day of the week (Table 2) and digit preference in BP became less common during the working week (aOR for Friday vs. Monday=0.87 [0.85-0.90]). As number of days into a hospital admission increased, temperatures were more likely to be recorded as 36.0°C in hospitals A-C with more marked increases at hospital C, but less likely in hospital B (Figure 2D). For BP, digit preference was relatively stable by day of admission and between hospitals, except at hospital B where it increased during hospital stay.

### Variation by specialty

Temperature readings of 36.0°C and BP readings ending in zero were generally more common in medical vs. surgical specialities (Figure 3). Exceptions included nephrology where value preference was less common for both temperature and BP, haematology and oncology where temperatures of 36.0°C were less commonly recorded and cardiology where BP digit preference was less common. Additionally, temperature readings of 36.0°C were more common in specialist surgery (ear, nose and throat surgery; plastic surgery; maxillofacial surgery; ophthalmology) and BP digit preference in haematology and oncology.

**Figure 3.**
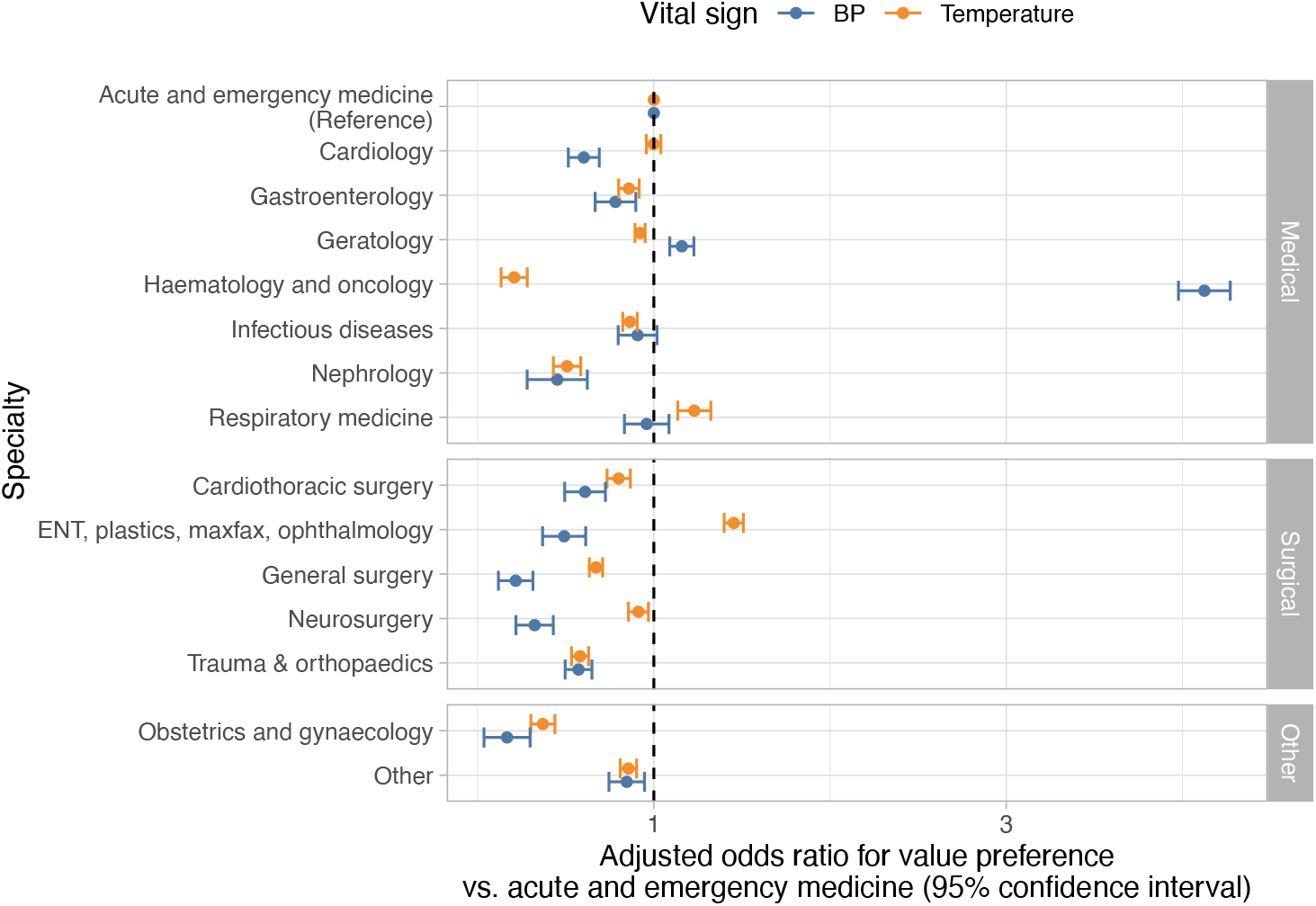
Relationship between specialty and temperature readings of 36.0°C and blood pressure readings ending in zero. ENT, ear, nose and throat; plastics, plastic surgery; maxfax, maxillofacial surgery; BP, blood pressure.

### Effect of previous abnormal measurements

A total of 4,037,232 sets of observations had a previous measurement from the same patient within ≤36 hours. Temperature readings of 36.0°C were less frequent following an abnormal prior temperature measurement, 20,272/351,619 (5.8%), than following a normal prior measurement, 587,856/3,685,613 (16.0%). Given it may take time for temperature to normalise it would not be expected that those with a previously abnormal temperature would have the same proportion of true temperatures of 36.0°C as the overall hospital population (estimated above as 4.9%). However even allowing for this, preference for recording a temperature of 36.0°C was more common with a normal prior measurement. In an analysis, adjusted for the same factors as in Table 1, a previous abnormal temperature reduced the odds of a recording of 36.0°C (aOR=0.35, [95%CI 0.34-0.35]). Similarly, 24,394/1,291,295 (1.9%) of BP observations had SBP and DBP both ending in zero after an abnormal SBP or DBP, compared to 66,253/2,745,937 (2.4%) without a prior abnormal reading (aOR=0.87 [0.85-0.89]).

## Discussion

In this analysis of records from a large UK teaching hospital group, we show preference for specific values or digits in vital sign records in EHRs. Our findings have implications for patient management, quality improvement initiatives and for research conducted using EHRs.

A key question is whether value preferences indicate simply convenience rounding on transcribing values or whether they are also a marker for incompletely observed observations. Potentially favouring the latter, we observed differences in the relative frequency of value preferences, with temperature recordings of 36.0°C three times more common than expected based on the remainder of the temperature distribution. It is also possible that given 36.0°C was the lower bound of the normal range in the hospitals’ early warning score, that implausibly low readings, e.g., when thermometers were mis-calibrated were recorded as 36.0°C. In contrast, BP readings ending in zero were twice as common, but HR readings ending in zero only around 20% above expected. This may reflect the additional burden of recording a patient’s temperature, requiring a second piece of equipment that may occasionally be missing or broken. Conversely it is possible that a greater proportion of all temperature values are rounded to a single value.

We found differences between hospital specialties, even after adjustment for other factors. Generally surgical specialities recorded vital signs with greater precision. However, the prevalence of value preferences also potentially reflects the culture within a speciality, where greater importance may be placed on measured values, e.g. in nephrology, or on specific vital signs, e.g. temperature in neutropenic and other immunosuppressed patients in haematology and oncology or BP in cardiology. We also found marked differences in temperature measurement between the four hospitals in the organisation, even following adjustment for the specialties present. This may reflect systemic factors, e.g., staffing levels and the importance placed on vital signs may vary by setting. In higher acuity settings, reliance on vital signs for treatment escalation could increase vital sign fidelity compared to less acute settings focused on rehabilitation. Although patients are admitted to acute medicine as an emergency, the increased digit preference seen in this specialty may reflect that for many longer staying patients rehabilitation and provision of social care are the dominant issues for much of each admission. In keeping with this, for both acute hospitals, A and C, we found that temperatures were more likely to be recorded as 36.0°C as length of stay increased. We also found that normal prior measurements were more likely to be followed by digital preference in subsequent observations, with previous abnormal measurements being associated with greater accuracy in subsequent observations.

Older patients and male patients were more likely to have vital sign recordings with value preferences. Further work is required to better understand the reasons for this. The reasons for variation in temperature recoding by age may reflect differences in the acuity of patients and associated culture around vital sign measurement, the relative importance placed on curative treatment vs. patient comfort, and physical barriers to temperature measurement including patient agitation. There were no systematic differences by ethnicity across both vital sign modalities.

Changes over time suggest institution-wide improvement is possible, with increased precision of both temperature and BP seen during the study. The study builds on previous studies of vital sign recording quality,^24^ and highlights that institutions may wish to monitor vital sign recording to identify areas of the hospital or patient groups where specific interventions to improve quality may be required.

Multiple variables representing the timing of measurements were investigated. Temperature appeared to be most likely to be recorded accurately in the afternoon or evening. Conversely BP measurements appeared most likely to be rounded in the evening. Generally, vital sign precision was greatest around the time of hospital admission, likely reflecting that patients are most unwell when first presenting to hospital and so vital signs are performed and recorded carefully and in full.

Digit preference is a well described phenomenon.^19,20^ However, particularly for temperature measurement, the question that arises from our findings is; if a vital sign is more difficult to measure for some reason then why does current culture potentially favour documenting an inaccurate reading instead of leaving it missing. There may be explicit or implied pressure to always record a complete set of vital signs (although ∼20% of observations in our study were excluded because of missing one or more vital signs), or it may be that recording an observation as unavailable may be more onerous and require entering a justification. There may also be disincentives to recording abnormal values if this requires escalation of care and additional action, but this would be expected to potentially impact all values rather than just those showing value preference.

Limitations of our study include that it is based on a single organisation and data entry system for recording vital signs. Further studies are required to confirm if our findings are replicated more widely. We did not investigate more granular variation in vital sign recording by hospital ward or individual staff member, the latter as the identity of the healthcare worker recording the vital signs was not available in our data extract. We also did not investigate the downstream consequences of vital sign value preferences, and this could be looked at in future work, e.g. considering associations with length of stay or mortality, although care would be required to avoid reverse causation where delays in discharge or a more palliative focus change value preferences.

Our study provides evidence that vital sign measurement displays value preference to a such a degree that it could affect conclusions based on unadjusted vital sign data, in both clinical and research settings. We show that hospital, speciality, admission stage and patient age all have important impacts on the accuracy of vital signs. Changes over time in our hospital suggest improvements in accuracy are possible. Ultimately fully connected systems that do not require re-entry of vital signs into the EHR are likely to address many of the issues identified. Work with institutions and individuals is required to fully elucidate and understand the mechanisms behind values preference on a systems, patient and clinician level. In the meantime, clinicians and researchers need to be aware that vital signs may not always be accurately documented, and to make appropriate allowances and adjustments for this in observational analyses using these factors as outcomes or exposures.

## Data Availability

The data analysed are not publicly available as they contain personal data but are available from the Infections in Oxfordshire Research Database (https://oxfordbrc.nihr.ac.uk/research-themes-overview/antimicrobial-resistance-and-modernising-microbiology/infections-in-oxfordshire-research-database-iord/), subject to an application and research proposal meeting on the ethical and governance requirements of the Database.

## Acknowledgements

This work uses data provided by patients and collected by the UK’s National Health Service as part of their care and support. We thank all the people of Oxfordshire who contribute to the Infections in Oxfordshire Research Database. Research Database Team: L Butcher, H Boseley, C Crichton, DW Crook, DW Eyre, O Freeman, J Gearing (community), R Harrington, K Jeffery, M Landray, A Pal, TEA Peto, TP Quan, J Robinson (community), J Sellors, B Shine, AS Walker, D Waller. Patient and Public Panel: G Blower, C Mancey, P McLoughlin, B Nichols.

## Transparency declarations

DWE declares lecture fees from Gilead outside the submitted work. No other author has a conflict of interest to declare.

## Supplementary Figure

**Figure S1.**
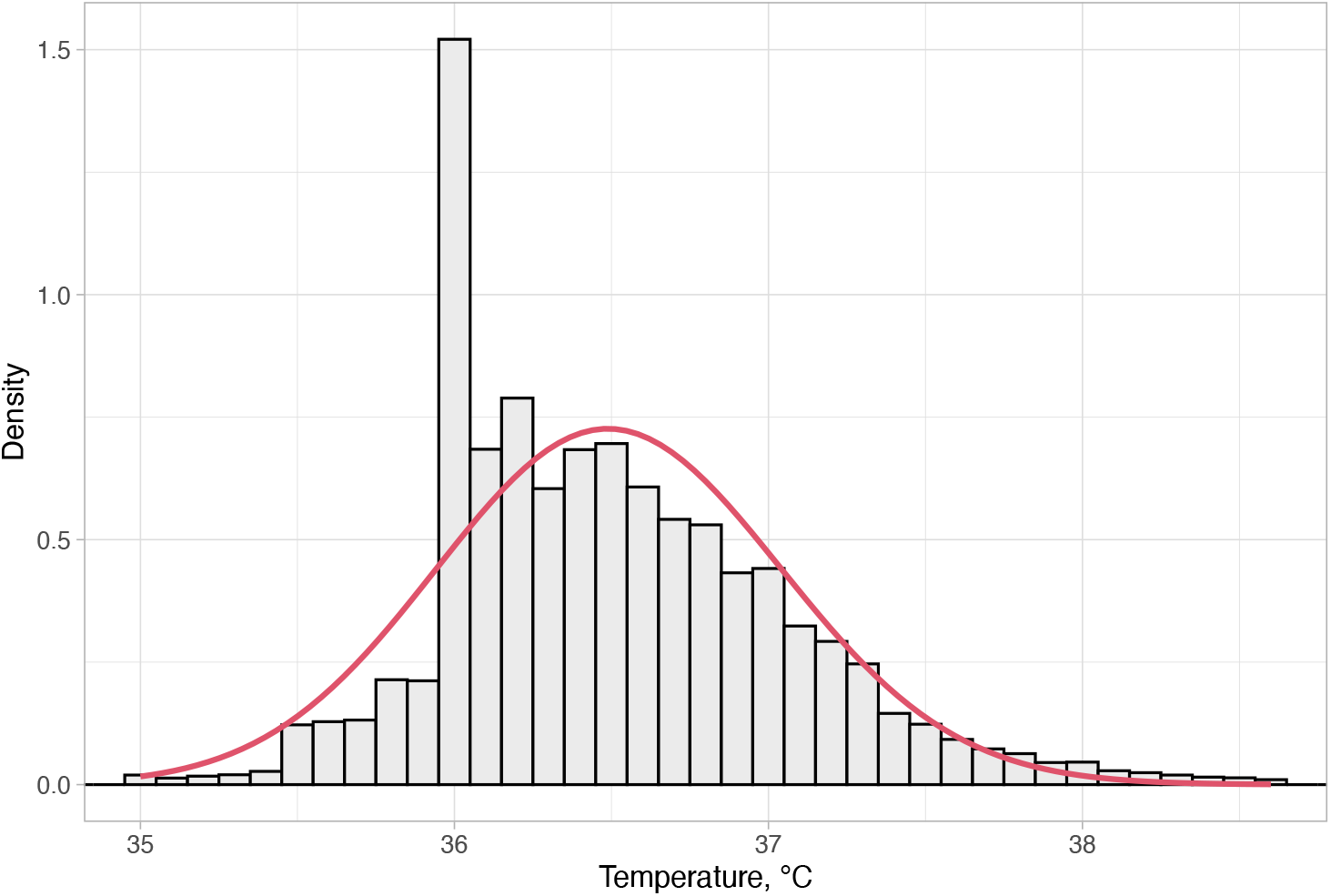
Observed and fitted distribution of temperature measurements. The red line indicates a gamma distribution fitted to the data. Based on the gamma distribution (with parameters: shape=4416, rate=121) 4.9% of values are expected to lie between 35.95°C and 36.05°C.

